# COVID-19 Case Age Distribution: Correction for Differential Testing by Age

**DOI:** 10.1101/2020.09.15.20193862

**Authors:** David N. Fisman, Amy L. Greer, Michael Hillmer, Sheila F. O’Brien, Steven J. Drews, Ashleigh R. Tuite

## Abstract

**Background:** SARS-CoV-2 is a novel pathogen and is currently the cause of a global pandemic. Despite expected universal susceptibility to a novel pathogen, the pandemic to date has been characterized by higher observed incidence in the oldest individuals and lower incidence in children and adolescents. Differential testing by age group may explain some of these observed differences, but datasets linking case counts to public health testing volumes are uncommon.

**Methods:** We used data from Ontario, Canada. Case data were obtained from Ontario’s provincial line, while testing data were obtained from an information system with complete SARS-CoV-2 testing data for public, hospital, and private laboratories. Demographic and temporal patterns in reported case incidence, testing rates, and test positivity were explored using negative binomial regression models. Standardized morbidity and testing ratios (SMR, STR), and standardized test positivity (STP) were calculated by dividing age- and sex-specific rates by overall rates; demographic and temporal patterns in standardized ratios were explored using meta-regression. Testing adjusted SMR were estimated using linear regression models.

**Results:** Observed disease incidence and testing rates were highest in oldest individuals and markedly lower in those aged < 20. Temporal trends in disease incidence and testing were observed, but standardizing morbidity and testing ratios eliminated temporal trends (i.e., relative patterns by age and sex remained identical regardless of epidemic phase). After adjustment for testing frequency, SMR were lowest in children and adults aged 70 and older, approximately the same in adolescents as in the population as a whole and elevated in young adults (aged 20-29 years), providing a markedly different picture of the epidemic than seen with crude SMR or case-based incidence. Test-adjusted SMR were validated using seroprevalence data (Pearson correlation coefficient 0.82, P = 0.04).

**Conclusions:** Surveillance for SARS-CoV-2 infection is typically performed using only test-positive case data, without adjustment for testing frequency. Older adults are tested more frequently, likely due to increased disease severity, while children are under-tested. Adjustment for testing frequency results in a very different picture of SARS-CoV-2 infection risk by age, one that is consistent with estimates obtained through serological testing.

## Background

The COVID-19 pandemic has a number of unusual features which have created controversy around optimal control strategies. One such unexpected feature is the apparent low incidence of infection by SARS-CoV-2 in children and adolescents (1, 2). Indeed, early in the pandemic there was a question of whether children might lack susceptibility to infection by SARS-CoV-2, though even an early report noted an asymptomatic child with pulmonary infection in association with a family cluster (3). A subsequent study in the Chinese city of Shenzhen demonstrated that infection in children is not rare (4). Early studies conducted under strict public health interventions found less evidence of active infection and seropositivity in children than older adults (5).

The failure to recognize pediatric infection early in the pandemic may have been due to the relative rarity of severe illness in younger people (2, 6), with resultant decreased testing rates. While severe illness and even death due to COVID-19 infection have been reported in children less than 10?, such occurrences are notably uncommon (6-9). In the Canadian province of Ontario, early challenges with laboratory testing for SARS-CoV-2 led to limited testing in those without severe symptoms; furthermore, the invasive nature of nasopharyngeal sampling for PCR testing makes it an unappealing modality for use in children.

The universal susceptibility to a novel disease in the context of a pandemic is expected to result in attack rates that are proportional to contact rates in a given age group. As children have the highest contact rates in society under normal circumstances one might expect attack rates in this age group to be higher rather than lower than those seen in the population as a whole (10). We postulated that the apparent decreased incidence of SARS-CoV-2 infection in children might reflect lower rates of testing in this age group, rather than any true biological difference in susceptibility. Our objective was to evaluate whether differences in COVID-19 incidence between children and adults in Ontario are accounted for by differences in testing. Our exploration of this question is facilitated by the existence of a single master record of all COVID-19 PCR tests completed in the province, a single master line list for all COVID-19 cases, in this jurisdiction, and aggregated blood donor serology data from Ontario collected by Canadian Blood Services.

## Methods

Ontario is Canada’s most populous province, with a current population of 14.7 million (11). The province identified imported COVID-19 cases from China, and Iran, in January and February 2020 (12); local epidemic spread of SARS-CoV-2 has been evident since late February, 2020 (13). Each of Ontario’s 34 public health units is responsible for local case investigation and uploading of case information into the iPHIS data system, which is used for surveillance and case management of notifiable diseases in the province (14). Ontario’s case definition for a confirmed case of SARS-CoV-2 requires a positive laboratory test using a validated nucleic acid amplification test, including real-time PCR and nucleic acid sequencing (15). Data were available on case age by 10-year intervals and sex, as well as date. We defined “children” as those aged 0-10, and “adolescents” as those aged 10-19.

Testing volumes were obtained from the Ontario Laboratory Information System (OLIS), which includes testing and reporting dates for all PCR tests performed in the province (16). While most testing for SARS-CoV-2 is performed in the province’s public health laboratory system, OLIS also contains records of testing performed at hospital and private laboratories, which have been contributing to testing since April 2020 in an effort to increase the province’s test capacity and reduce turn-around times.

Testing data and case data are not linkable at the individual level, but daily case counts and test counts, by age and sex, were linked by report date, a field common to both the OLIS and iPHIS datasets. Our analysis was restricted to the time period between March 1, 2020 and July 26, 2020, with the earlier timepoint representing the start of the first month during which community transmission of SARS-CoV-2 was clearly occuring in Ontario (13, 17). Age- and sex-specific populations derived from Statistics Canada were used for estimation of disease and testing cumulative incidence (18). Cumulative incidence estimates were annualized by dividing populations by the time period under study to convert them to “person-years at risk”.

We explored rates of testing, diagnosis and per-test positivity using negative binomial regression models with populations as offsets (for the former two analyses) or test volumes as offsets (for the latter). We used 10-year age categories. Age categories were treated as (0,1) indicator variables, with age 50-59 used as a referent; we explored changing test rates by month, with March 2020 used as a referent. Standardized morbidity, testing and test positivity ratios overall (SMR, STR, and STP, respectively), and by month, were estimated by calculating cumulative incidence of disease or testing, or positivity per test, for the population as a whole, and then by age and sex subgroups. Ratios were then estimated by dividing subgroup specific estimates by estimates for the population as a whole.

Confidence intervals were calculated using estimates for standard error of ln(ratios), estimated as

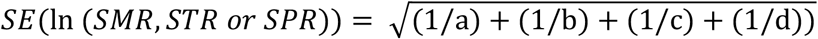

for ln(ratios), where a is the test or case count in the population subgroup, b is the population (or in the case of positivity, the test count) in the population subgroup, c is (overall test or case count – a), and d is (overall population or test count – b). By convention SMR would be multiplied by 100, but we avoided this multiplier for methodological reasons described below. Differences in SMR, STR, and STP by age group and gender, and over time, were explored through construction of meta-regression models weighted using standard error estimates as described above.

We postulated that low SMR in children might be explained by lower testing rates. To estimate “test adjusted” estimates of SMR, we created age-specific linear regression models, with log(STR) used as the independent variable, and SMR the dependent variable. As log(STR) is zero when testing in a given age group is equivalent to the overall population test rate, the intercepts from these models provide an estimate of expected SMR, if the age group in question were tested at the same rate as the population overall. We validated this approach by comparing test-adjusted SMR derived in this way from SMR derived from Ontario seroprevalence estimates derived from blood donor populations, though these estimates were only available for individuals aged 17 and older.

Briefly, data represented residual donor plasma specimens tested at Canadian Blood Services using the Abbott Architect SARS-CoV-2 IgG (Abbott, Chicago, IL, USA)

Test-adjusted SMR were then used to generate estimates of test-adjusted incidence, which would be perceived if all age groups were tested at the same rate as the most frequently tested (oldest) age group. If I_iTmax_ is incidence in the maximally tested i^th^ age group, and I_o_ is incidence in the population overall, then I_o_ in a maximally tested population is I_iTmax_/SMR_i_. For any other i^th^ age group, test-adjusted incidence in the face of maximal testing is then simply SMR_i_ multiplied by I_o_.

All analyses were performed using Stata SE version 15.0 (College Station, Texas). The study received ethics approval from the Research Ethics Board at the University of Toronto.

## Results

Between March 1 and July 26, 2020, 38,405 cases of COVID-19 were diagnosed in Ontario, while unique individuals were tested for COVID-19. Annualized testing rates increased 10-fold from 3060 tests per 100,000 population in March to 33,162 per 100,000 in July (**Figure 1**); a total of 1.33 million tests were performed during this time period. At the same time annualized crude disease incidence declined from a peak of 1287 cases per 100,000 in April 2020 to 260 per 100,000 by July. In negative binomial models, both case and test rates were lowest in those aged <20, and highest in those aged over 60. Males were less likely to be tested than females, but per-test positivity was higher in males than females. Per-test positivity declined from April to July (**Table 1**).

**Figure 1.**
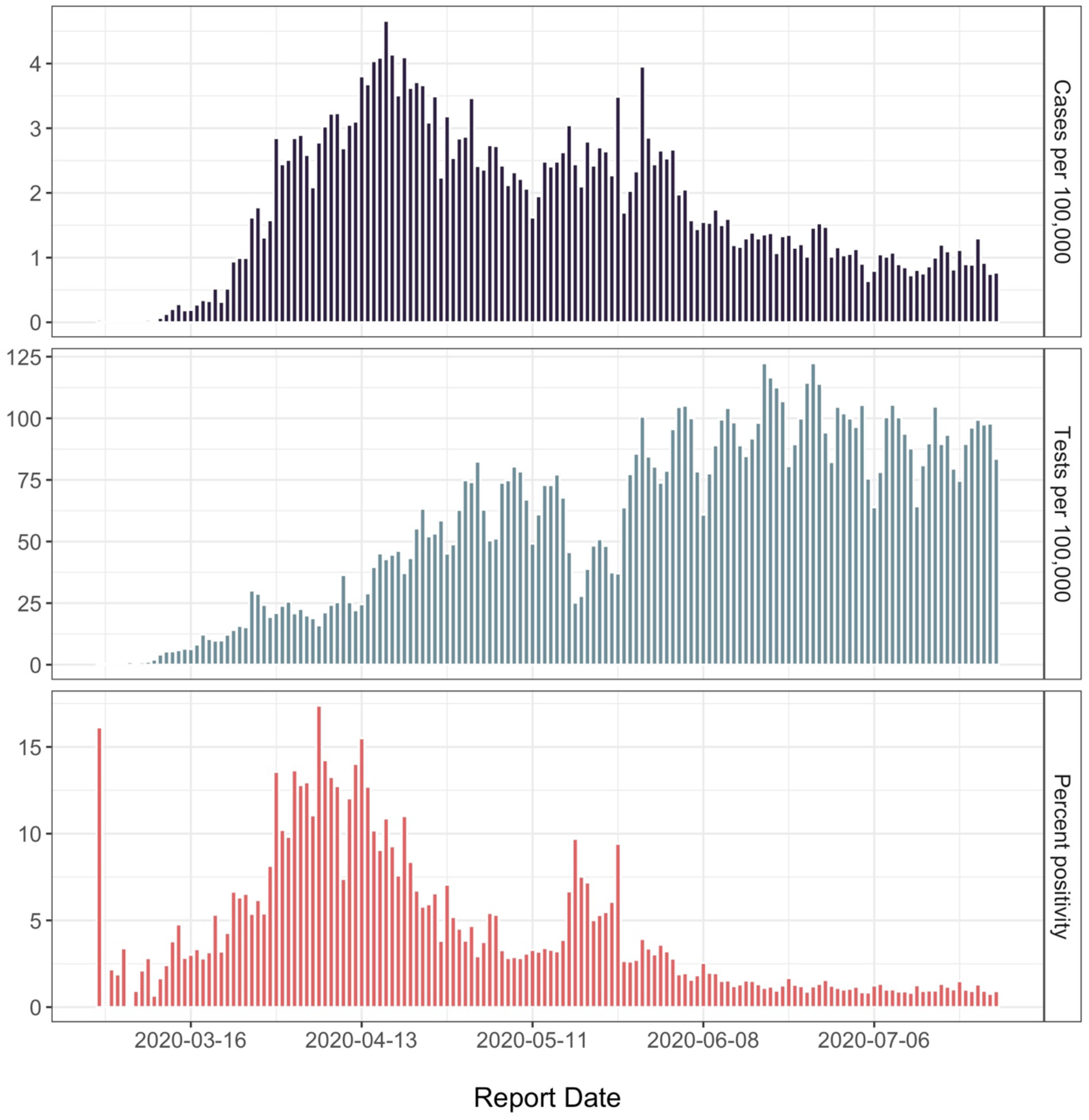
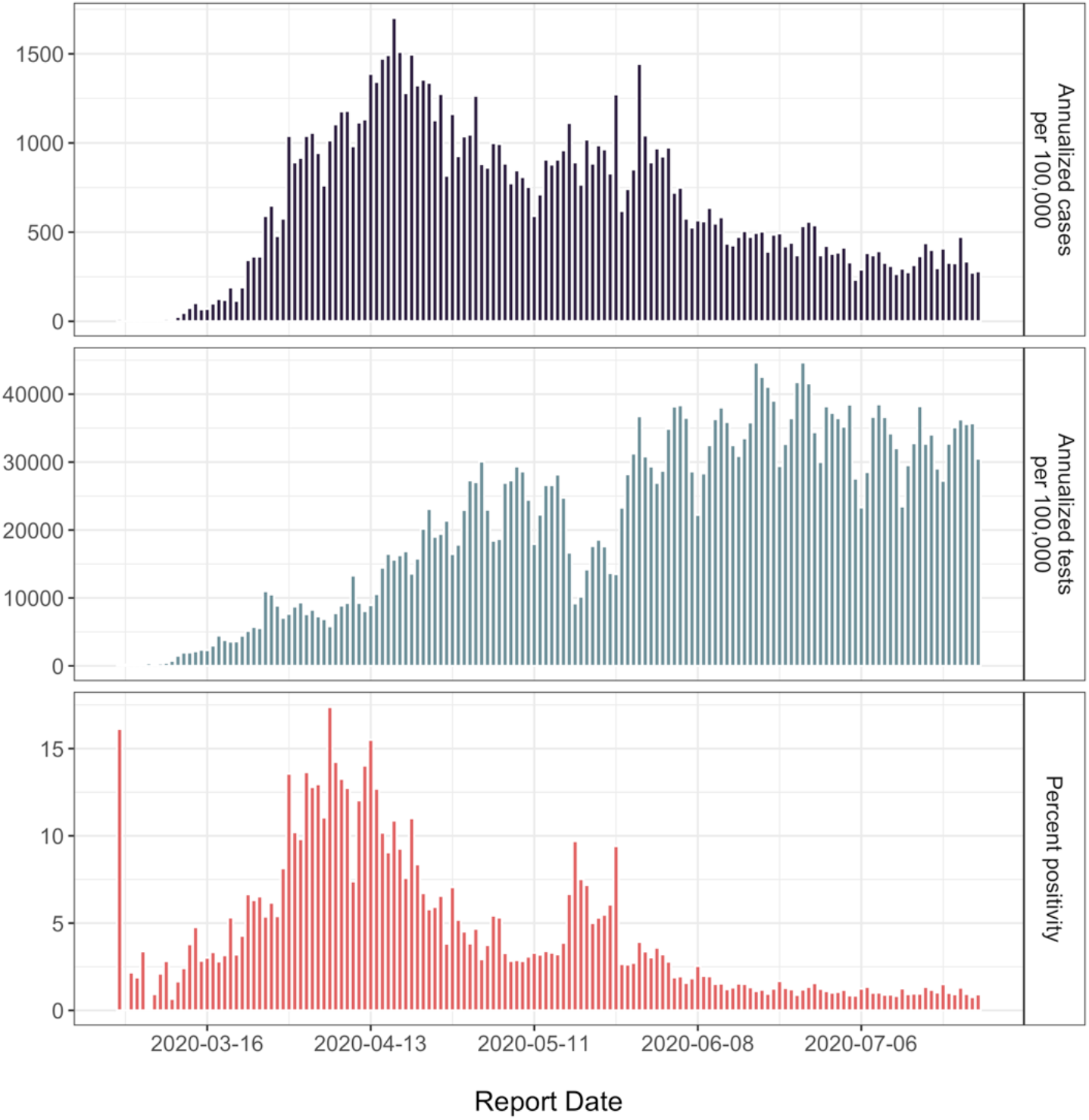
Epidemiology of the SARS-CoV-2 Pandemic in Ontario, Canada, March 1-July 26, 2020. The panels demonstrate decreasing infection incidence over time (top panel), accompanied by increasing use of testing resources over the same time period (middle panel). Per-test positivity peaked in April, 2020 (bottom panel).

**Table 1.**
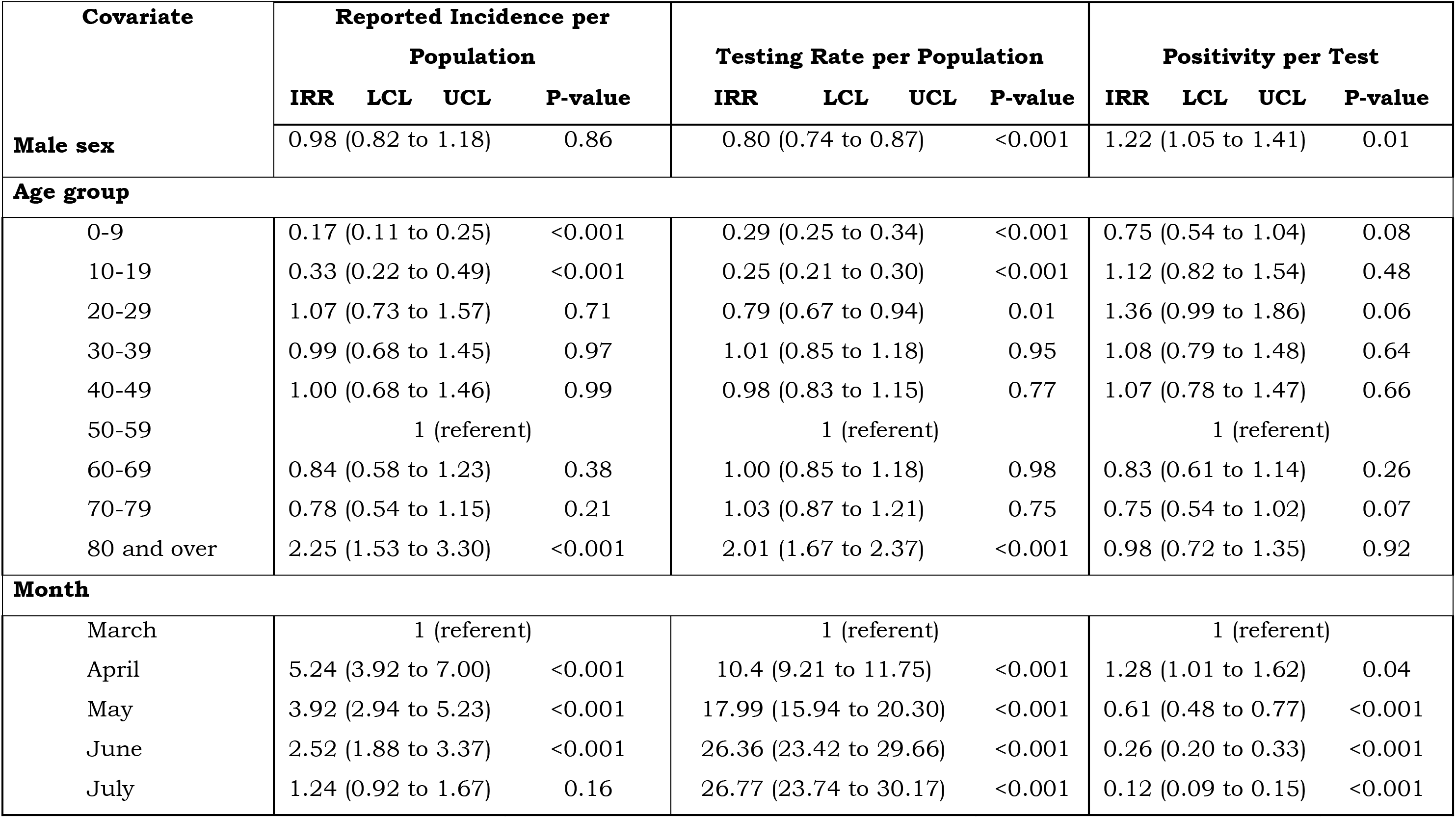
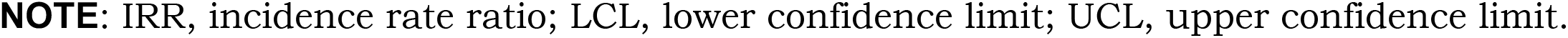
Negative Binomial Models of COVID-19 Disease Incidence, Testing Incidence, and Positivity per Test, Ontario, Canada.

| Covariate | Reported Incidence per Population |  |  |  | Testing Rate per Population |  |  |  | Positivity per Test |  |  |  |
| --- | --- | --- | --- | --- | --- | --- | --- | --- | --- | --- | --- | --- |
|  | IRR | LCL | UCL | P-value | IRR | LCL | UCL | P-value | IRR | LCL | UCL | P-value |
| Male sex | 0.98 (0.82 to 1.18) |  |  | 0.86 | 0.80 (0.74 to 0.87) |  |  | <0.001 | 1.22 (1.05 to 1.41) |  |  | 0.01 |
| Age group |  |  |  |  |  |  |  |  |  |  |  |  |
| 0-9 | 0.17 (0.11 to 0.25) |  |  | <0.001 | 0.29 (0.25 to 0.34) |  |  | <0.001 | 0.75 (0.54 to 1.04) |  |  | 0.08 |
| 10-19 | 0.33 (0.22 to 0.49) |  |  | <0.001 | 0.25 (0.21 to 0.30) |  |  | <0.001 | 1.12 (0.82 to 1.54) |  |  | 0.48 |
| 20-29 | 1.07 (0.73 to 1.57) |  |  | 0.71 | 0.79 (0.67 to 0.94) |  |  | 0.01 | 1.36 (0.99 to 1.86) |  |  | 0.06 |
| 30-39 | 0.99 (0.68 to 1.45) |  |  | 0.97 | 1.01 (0.85 to 1.18) |  |  | 0.95 | 1.08 (0.79 to 1.48) |  |  | 0.64 |
| 40-49 | 1.00 (0.68 to 1.46) |  |  | 0.99 | 0.98 (0.83 to 1.15) |  |  | 0.77 | 1.07 (0.78 to 1.47) |  |  | 0.66 |
| 50-59 | 1 (referent) |  |  |  | 1 (referent) |  |  |  | 1 (referent) |  |  |  |
| 60-69 | 0.84 (0.58 to 1.23) |  |  | 0.38 | 1.00 (0.85 to 1.18) |  |  | 0.98 | 0.83 (0.61 to 1.14) |  |  | 0.26 |
| 70-79 | 0.78 (0.54 to 1.15) |  |  | 0.21 | 1.03 (0.87 to 1.21) |  |  | 0.75 | 0.75 (0.54 to 1.02) |  |  | 0.07 |
| 80 and over | 2.25 (1.53 to 3.30) |  |  | <0.001 | 2.01 (1.67 to 2.37) |  |  | <0.001 | 0.98 (0.72 to 1.35) |  |  | 0.92 |
| Month |  |  |  |  |  |  |  |  |  |  |  |  |
| March | 1 (referent) |  |  |  | 1 (referent) |  |  |  | 1 (referent) |  |  |  |
| April | 5.24 (3.92 to 7.00) |  |  | <0.001 | 10.4 (9.21 to 11.75) |  |  | <0.001 | 1.28 (1.01 to 1.62) |  |  | 0.04 |
| May | 3.92 (2.94 to 5.23) |  |  | <0.001 | 17.99 (15.94 to 20.30) |  |  | <0.001 | 0.61 (0.48 to 0.77) |  |  | <0.001 |
| June | 2.52 (1.88 to 3.37) |  |  | <0.001 | 26.36 (23.42 to 29.66) |  |  | <0.001 | 0.26 (0.20 to 0.33) |  |  | <0.001 |
| July | 1.24 (0.92 to 1.67) |  |  | 0.16 | 26.77 (23.74 to 30.17) |  |  | <0.001 | 0.12 (0.09 to 0.15) |  |  | <0.001 |

**NOTE:** IRR, incidence rate ratio; LCL, lower confidence limit; UCL, upper confidence limit.

**Table 2.**
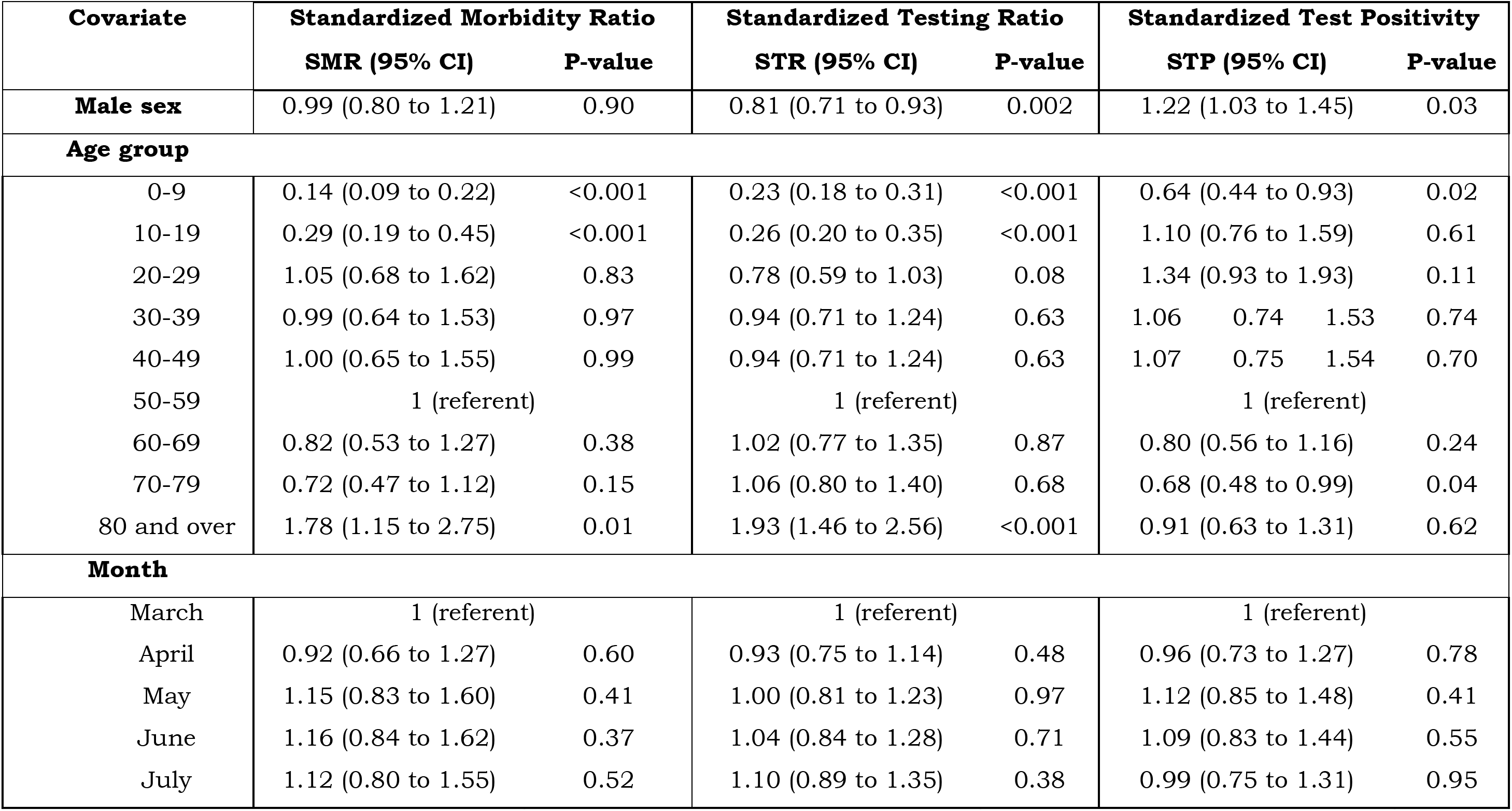
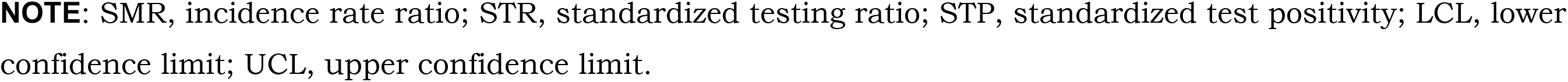
Meta-regression Models of Effects of Sex, Age and Month on Standardized Morbidity Ratios, Standardized Testing Ratios, and Standardized Test Positivity, Ontario, Canada

| **Covariate** | **Standardized Morbidity Ratio** |  | **Standardized Testing Ratio** |  | **Standardized Test Positivity** |  |
| --- | --- | --- | --- | --- | --- | --- |
|  | **SMR (95% CI)** | **P-value** | **STR (95% CI)** | **P-value** | **STP (95% CI)** | **P-value** |
| **Male sex** | 0.99 (0.80 to 1.21) | 0.90 | 0.81 (0.71 to 0.93) | 0.002 | 1.22 (1.03 to 1.45) | 0.03 |
| **Age group** |  |  |  |  |  |  |
| 0-9 | 0.14 (0.09 to 0.22) | <0.001 | 0.23 (0.18 to 0.31) | <0.001 | 0.64 (0.44 to 0.93) | 0.02 |
| 10-19 | 0.29 (0.19 to 0.45) | <0.001 | 0.26 (0.20 to 0.35) | <0.001 | 1.10 (0.76 to 1.59) | 0.61 |
| 20-29 | 1.05 (0.68 to 1.62) | 0.83 | 0.78 (0.59 to 1.03) | 0.08 | 1.34 (0.93 to 1.93) | 0.11 |
| 30-39 | 0.99 (0.64 to 1.53) | 0.97 | 0.94 (0.71 to 1.24) | 0.63 | 1.06 0.74 1.53 | 0.74 |
| 40-49 | 1.00 (0.65 to 1.55) | 0.99 | 0.94 (0.71 to 1.24) | 0.63 | 1.07 0.75 1.54 | 0.70 |
| 50-59 | 1 (referent) |  | 1 (referent) |  | 1 (referent) |  |
| 60-69 | 0.82 (0.53 to 1.27) | 0.38 | 1.02 (0.77 to 1.35) | 0.87 | 0.80 (0.56 to 1.16) | 0.24 |
| 70-79 | 0.72 (0.47 to 1.12) | 0.15 | 1.06 (0.80 to 1.40) | 0.68 | 0.68 (0.48 to 0.99) | 0.04 |
| 80 and over | 1.78 (1.15 to 2.75) | 0.01 | 1.93 (1.46 to 2.56) | <0.001 | 0.91 (0.63 to 1.31) | 0.62 |
| **Month** |  |  |  |  |  |  |
| March | 1 (referent) |  | 1 (referent) |  | 1 (referent) |  |
| April | 0.92 (0.66 to 1.27) | 0.60 | 0.93 (0.75 to 1.14) | 0.48 | 0.96 (0.73 to 1.27) | 0.78 |
| May | 1.15 (0.83 to 1.60) | 0.41 | 1.00 (0.81 to 1.23) | 0.97 | 1.12 (0.85 to 1.48) | 0.41 |
| June | 1.16 (0.84 to 1.62) | 0.37 | 1.04 (0.84 to 1.28) | 0.71 | 1.09 (0.83 to 1.44) | 0.55 |
| July | 1.12 (0.80 to 1.55) | 0.52 | 1.10 (0.89 to 1.35) | 0.38 | 0.99 (0.75 to 1.31) | 0.95 |

**NOTE:** SMR, incidence rate ratio; STR, standardized testing ratio; STP, standardized test positivity; LCL, lower confidence limit; UCL, upper confidence limit.

Both standardized test ratios and standardized morbidity ratios (**Figure 2**) were decreased in children and greater than 1 in older individuals, consistent with both increased case identification and increased testing in the older population. Standardized test positivity was lowest in children, but above 1 in young and middle-aged males. In meta-regression models evaluating the effects of age, sex and month on SMR and STR, age and gender effects were similar to those seen in negative binomial models but did not demonstrate time trends. Meta-regression models for STP demonstrated decreased positivity in the youngest age group, and elevated positivity in males; again, no differences in standardized ratios were seen by month.

**Figure 2.**
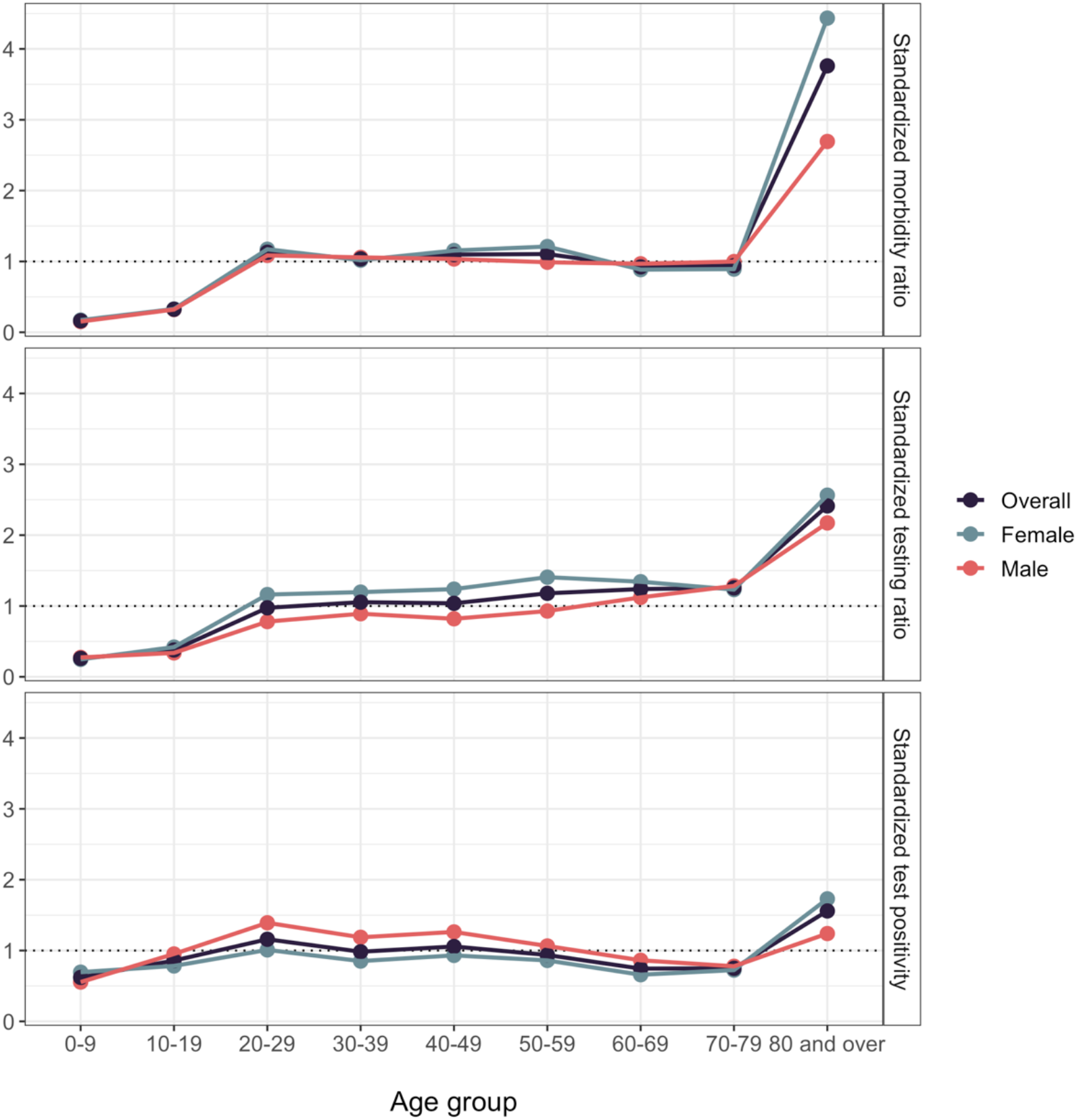
Standardized Disease and Testing Ratios During the SARS-CoV-2 Pandemic in Ontario, Canada, March 1-July 26, 2020. Ratios are calculated by dividing disease incidence (top), test incidence (middle), or test positivity (bottom) within a given age- and sex-group by incidence or test positivity in the population overall. Circles represent point estimates. Ratio of 1 indicates that within-group incidence or positivity is equivalent to that in the population as a whole.

Intercepts from linear models regressing SMR on log(STR) (interpreted as SMR when testing in a given age group is equivalent to that in the population as a whole) are presented in **Table 3**. Notably, the test-adjusted SMR was < 1 in both children aged < 10, and in adults over age 70. Test-adjusted SMR was 0.90 (95% CI 0.81-0.99) in adolescents, and substantially greater than 1 (SMR = 1.22, 95% CI 1.15-1.30) in individuals aged 20 to 29. In those aged 20 and older, test-adjusted SMR was highly correlated with seroprevalence-derived SMR (**Table 3**, rho = 0.82, P = 0.04). Test-adjusted SMR was weakly correlated with STP (rho = 0.66, P = 0.08), and STP and seroprevalence-derived SMR were not correlated (rho = 0.30, P = 0.55) (**Figure 3**).

**Table 3.** Test-adjusted Standardized Morbidity Ratios, and Standardized Morbidity Ratios Derived from Canadian Blood Services Seroprevalence Estimates, Ontario, Canada

| Age Group | SMR (Test-adjusted) (95% CI) | SMR (Serology-based) (95% CI) |
| --- | --- | --- |
| 0-9 | 0.24 (0.16 to 0.32) | — |
| 10-19 | 0.90 (0.81 to 0.99) | — |
| 20-29 | 1.22 (1.15 to 1.30) | 1.47 (1.06 to 2.02) |
| 30-39 | 1.09 (1.01 to 1.16) | 0.83 (0.55 to 1.26) |
| 40-49 | 1.16 (1.06 to 1.27) | 0.72 (0.45 to 1.15) |
| 50-59 | 1.08 (0.99 to 1.16) | 1.11 (0.77 to 1.60) |
| 60-69 | 1.15 (0.92 to 1.38) | 1.04 (0.68 to 1.58) |
| 70+* | 0.67 (0.49 to 0.85) | 0.29 (0.09 to 0.92) |

**NOTE:** SMR, incidence rate ratio; LCL, lower confidence limit; UCL, upper confidence limit.
\*Oldest age groups combined for consistency with seroprevalence data.

**Figure 3.**
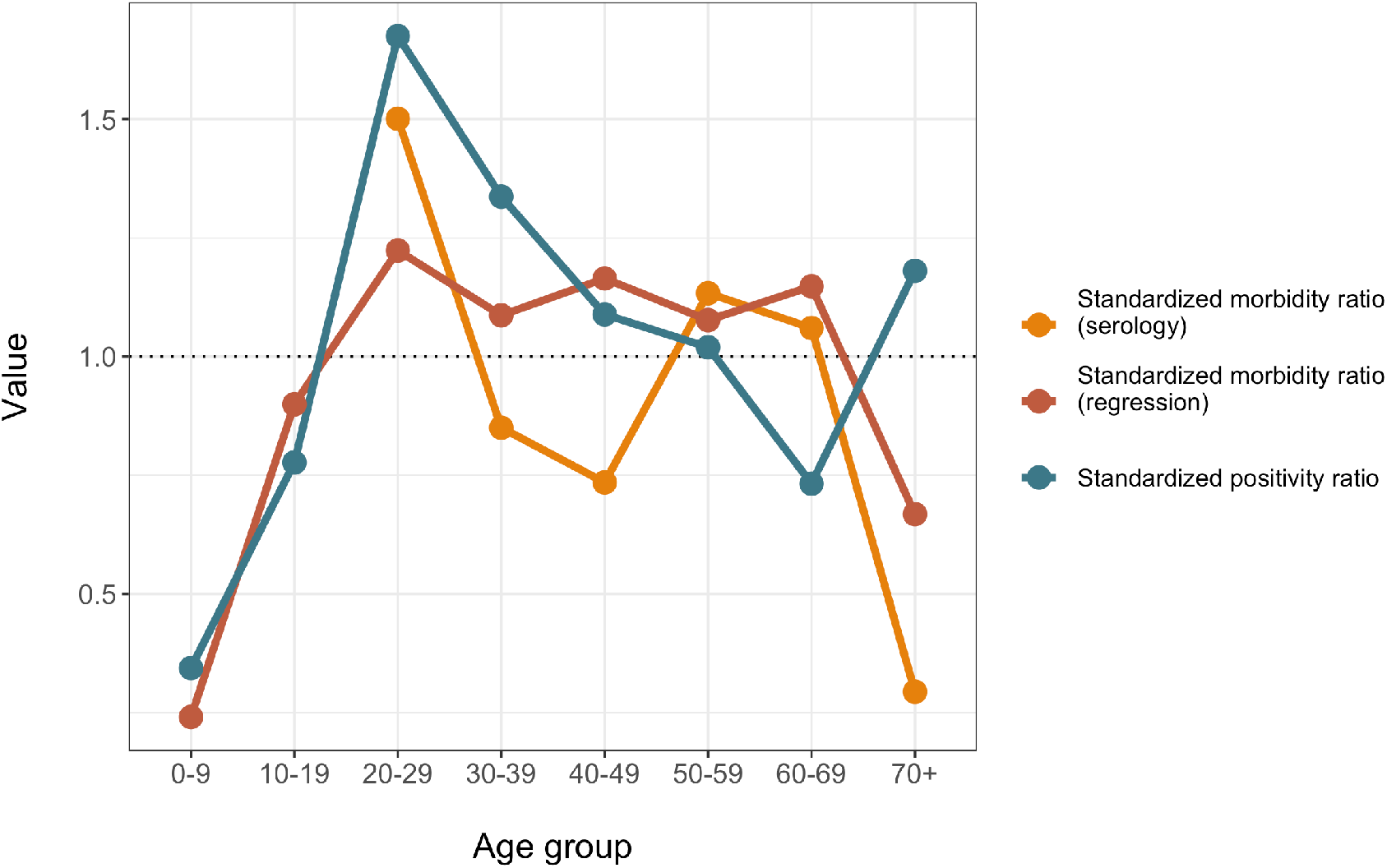
Comparison of Standardized Morbidity Ratios Derived via Test-Adjustment and Serological Testing, and Standardized Test Positivity. Test-adjusted standardized morbidity ratios (SMR) derived through regression (red) and from serological data (orange), and standardized test positivity (STP) (green) are plotted by age group. Test-adjusted SMR were highly correlated with seroprevalence-derived SMR, and weakly correlated with STP. Ratio of 1 indicates that within-group incidence or positivity is equivalent to that in the population as a whole.

When we assumed that maximal testing had been applied to those aged 70 and over, we estimated that the test-adjusted cumulative incidence in the Province as of July 26, 2020 was 1385 cases per 100,000 population, as compared to a crude cumulative incidence of 282 per 100,000 without test adjustment. Estimated testing-adjusted incidence was markedly higher than observed incidence in all age groups < 70 years (**Figure 4**). The fraction of cases identified, relative to expectation if all ages were tested with the same intensity as adults over 70, was 20%, almost identical to estimates derived using seroprevalence data (18%).

**Figure 4.**
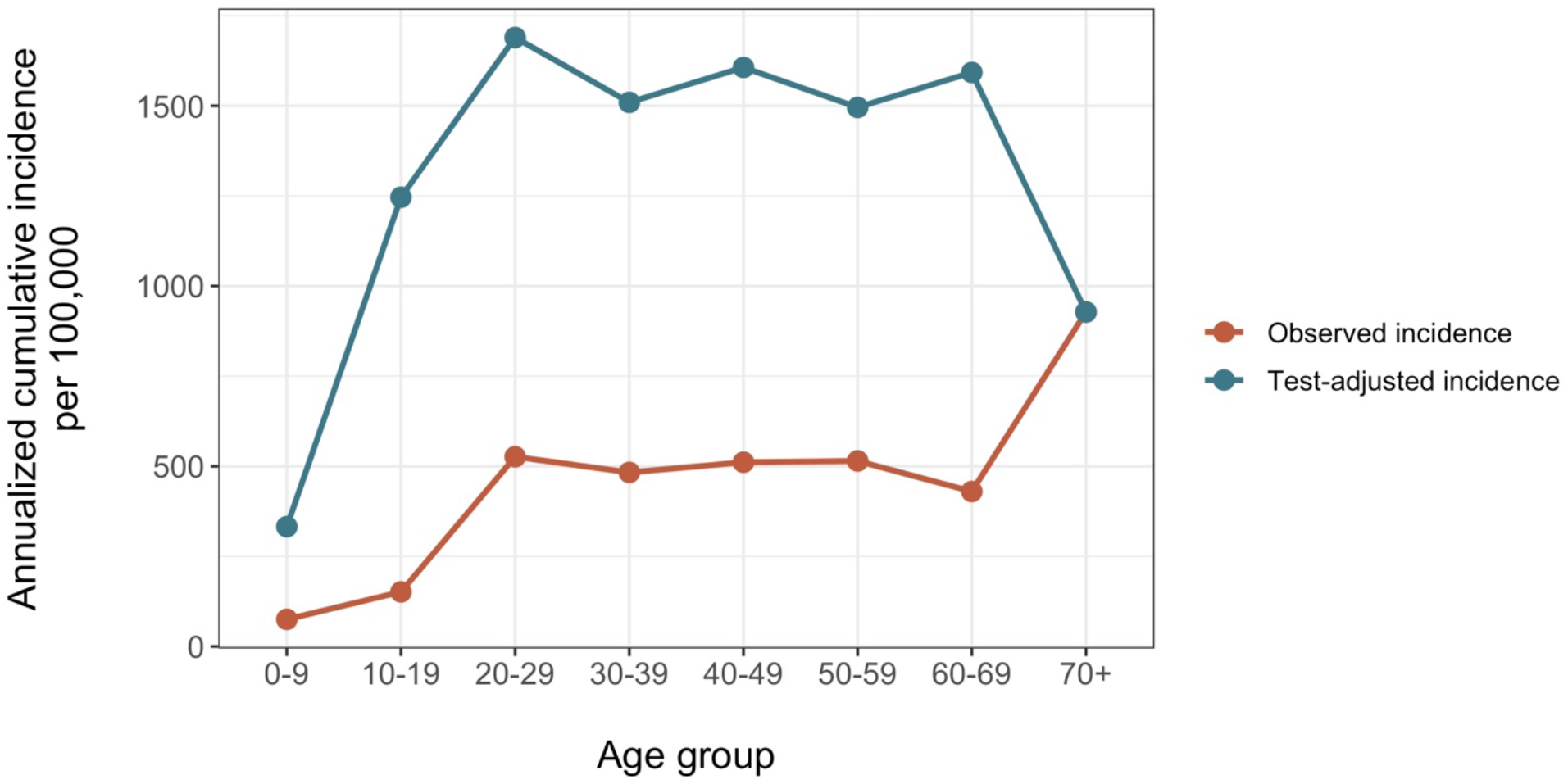
Observed and Test-Adjusted Estimates of Cumulative Incidence of SARS-CoV-2 Infection, Ontario, Canada. Observed cumulative incidence of SARS-CoV-2 infection in Ontario, Canada, by age, to July 26, 2020 (blue curve). Estimates were test-adjusted using standardized morbidity ratios as described in the text, under the assumption that maximal testing was performed in individuals aged 70 and over. The resultant test-adjusted estimates of cumulative incidence are plotted in the red curve.

## Discussion

The perceived epidemiology of reportable communicable diseases is often based exclusively on reports of test-positive cases, or cases meeting case definitions, without reference to how many individuals are tested or assessed as possible cases. However, the identification of cases generally depends on diagnostic testing, and differential testing volumes may dramatically alter how an epidemic is perceived. For most diseases of public health concern, surveillance systems do not incorporate test denominators, with influenza surveillance based on percent positivity of tests being a notable exception (19). We were able to evaluate both case counts, and test counts, for SARS-CoV-2 in a single large jurisdiction with a single testing authority. We found that standardized ratios (whether SMR, STR or STP) had several attractive properties: they reproduced estimates of relative risk that were remarkably similar to those derived using more conventional count-based regression models, they remained stable over time notwithstanding marked changes in disease risk over the several month periods, and they allowed us to estimate relative risk without having to arbitrarily designate a particular age group, sex or time period as a referent. The fact that standardized ratios remained fairly constant over time suggests that relative patterns of risk and testing in the population were stable regardless of epidemic stage.

We found that accounting for STR in estimating SMR resulted in a markedly different view of the epidemic. This method, which builds on earlier work by Grewelle and DeLeo for estimation of infection fatality ratio (20), requires few assumptions and should be readily implementable by local and regional public health authorities. Adjustment for test volume had marked, but opposite, effects on perceived disease risk at the extremes of age. In all age groups < 30 years, test-adjusted SMR was substantially higher than unadjusted SMR. In the oldest age groups, test-adjusted SMR declined substantially, relative to unadjusted SMR. While test-adjusted SMR in the youngest age group remained substantially below 1, it was nearly double the unadjusted value. These results, which we were able to partially validate using seroprevalence data from individuals older than 18 years of age, suggest that the high rates of reported COVID-19 in older adults are most likely due to increased testing due to increased disease severity (21); in fact, older adults may be at less risk of infection than younger individuals, possibly due to greater adherence with social distancing, masking and other protective behaviors (22). By contrast, adults aged 20-29 are at markedly higher risk of infection after adjustment for decreased testing frequency; this again likely reflects risk perceptions and lack of adherence to preventive measures (22), while adolescents and teens had test-adjusted estimates of risk similar to those in the population as a whole.

The finding that children 10 and older are infected at rates similar to the population as a whole, after adjustment for testing frequency, is consistent with expectations for a pandemic disease, in which high attack rates reflect initial universal susceptibility to disease. After adjusting for testing frequency, the SMR in children increased approximately 2-fold (from 0.14 to 0.24) but remained substantially lower than the population as a whole. While younger children have appeared less likely to be infected in both PCR-based population screening studies and serological surveys (5, 23, 24), we would caution against ascribing this apparent decrease in risk to biological or immunological mechanisms. Younger children are likely to have been more compliant with social distancing than adolescents with more autonomy; have been deprived of typical school-based contact networks; and may have atypical presentations of SARS-CoV-2 infection, such as gastrointestinal illness, such that children presenting with symptoms of true infection may have been systematically under-tested (25). In terms of implications of our findings for settings such as schools, we would thus caution against over-reliance on surveillance data that do not include testing of asymptomatic children, and children without typical respiratory symptoms, to draw conclusions about COVID-19 epidemiology in a given locale, as resulting estimates of prevalence may be misleading.

Our method of test-adjustment was validated using SMR derived from blood donor data, which unfortunately limits our ability to validate test-adjusted SMR estimates in children, who do not donate blood. Nonetheless, the close correlation between test-adjusted SMR and serology-based SMR provides a degree of confidence in the soundness of this method; in particular, both serology-based SMR and test-adjusted SMR show that disease incidence is in fact higher in young adults, and lower in the elderly, than case-based surveillance data would suggest. Furthermore, apparent attenuation of risk at the extremes of age is consistent with serological data from other centres (e.g., a recent seroprevalence study performed in Geneva, Switzerland (26)).

Our study is made possible by the unusual (in the North American context) transparency of health agencies in a single large North American jurisdiction, which allows linkage of both test and case data, including data on testing volumes for both cases and non-cases. This provides a unique opportunity to evaluate the degree to which testing is a driver of the perceived severity of an epidemic. However, our study is nonetheless subject to several limitations, most notably our inability to directly link individuals’ case data with the test dataset. Our ability to validate our test-adjustment method is also limited by lack of availability of pediatric serological data, and it should also be noted that our seroprevalence estimates are based on blood donor samples, and might be less representative of population patterns of infection than results from a purposive, population-based serosurvey. Lastly, our results reflect epidemiology at an early timepoint in a single, high-income, North American jurisdiction. Nonetheless, we are able to demonstrate that test-adjustment provides a markedly different view of SARS-CoV-2, one that is consistent with serological results rather than those derived from a traditional case-based surveillance approach. While the work presented here awaits validation in other settings, it potentially provides a simple, inexpensive approach to more nuanced estimation of infection risk, by age, in jurisdictions that currently lack seroprevalence data.

## Data Availability

Individual-level data are not available. For access to aggregate data please contact Dr. Fisman.

## Notes

The research was supported by a grant to DNF from the Canadians Institutes for Health Research (2019 COVID-19 rapid researching funding OV4-170360).

### Competing Interest Statement

The authors have declared no competing interest.

### Author Declarations

Approved by the Research Ethics Board, University of Toronto

